# Self-applied single-channel mastoid ExG for fully automated detection of REM sleep behaviour disorder

**DOI:** 10.64898/2026.08.14.26360475

**Authors:** Casper Skjærbæk, Andreas Tind Damgaard, Natasha Becker Bertelsen, Thea Pinholt Lillethorup, Jacob Horsager, Victoria Lowe, Naja Helt Andersen, Astrid Juhl Terkelsen, Marit Otto, David Bertram, Martin Rodemann, Simon Lind Kappel, Yousef Tabar, Michael Sommerauer, Per Borghammer, Preben Kidmose

## Abstract

Isolated REM sleep behaviour disorder (RBD) is the strongest prodromal marker of Parkinson’s disease (PD) and dementia with Lewy bodies, yet diagnosis requires video-polysomnography with assisted montage and expert scoring and does not scale to screening or trial enrichment. We developed a fully automated, self-applied system that detects RBD from a pair of electrodes placed behind the ears, with no manual scoring at any stage. A novel bipolar mastoid ExG derivation enables both sleep staging and quantification of REM sleep without atonia (RWA). A fine-tuned deep-learning 1-channel model staged sleep at a Cohen’s kappa of 0.65 in PD, iRBD and controls (κ = 0.73 for the 2-channel model). Automated mastoid RWA correlated strongly with expert chin SINBAR scoring (r = 0.82). In self-applied home recordings from 76 participants, the 1-channel system detected RBD with an AUC of 0.95 (sensitivity 94%, specificity 86%), reproduced on in-lab polysomnographies (AUC 0.93, n = 378). In RBD, between-night RWA variability warrants repeated nights for prognostic monitoring. The system provides a scalable tool for RBD detection and a continuous RWA measure for longitudinal studies of neurodegeneration.

## Introduction

Sleep disturbances are common in the α-synucleinopathies, including Parkinson’s disease (PD), dementia with Lewy bodies (DLB) and multiple system atrophy, even in the prodromal stage^1,2^. Their clinical relevance extends beyond sleep disruption, as these disorders are closely linked to REM sleep behaviour disorder (RBD), characterised by recurrent dream-enactment behaviour and loss of muscle atonia during REM sleep, termed REM sleep without atonia (RWA). When RBD occurs without manifest neurological disease, isolated RBD (iRBD) is the strongest clinically identifiable prodromal marker of PD^3–5^. In the largest multicentre cohort (n = 1,280), 73.5% of patients with iRBD phenoconverted to parkinsonism or dementia within 12 years (annual conversion rate 6.3%)^6^, exceeding 90% at longer follow-up^7^. This almost inevitable progression, but prolonged prodromal interval, makes isolated RBD an important population for observational research, biomarker development, and neuroprotective trials. With community-based screening more individuals could be identified, and at an earlier clinical stage than case-finding through specialist sleep centres^8^. Realising this opportunity requires objective identification methods that can extend beyond established referral pathways and be deployed at scale.

Detection of RBD remains important after PD onset, as patients with RBD (PD^RBD+^) carry greater cognitive and non-motor burden than those without RBD (PD^RBD-^)^9–12^, and RBD features in early PD predict faster motor and cognitive decline^13,14^. RBD also informs PD subtyping, including the body-first versus brain-first model and data-driven mild-motor-predominant versus diffuse-malignant phenotypes^13,15^. This has direct clinical relevance, as post hoc analyses of the recent PASADENA trial of prasinezumab in PD found slower motor progression in the diffuse-malignant subtype and in patients with symptoms of RBD despite a missed primary endpoint^16,17^. These findings need confirmation but illustrate why objective RBD status could support cohort characterisation, prognostic enrichment and trial stratification. Beyond binary status, quantitative RWA captures phenoconversion risk and may serve as a biomarker of treatment response ^18,19^, while reliable sleep architecture measurements could guide sleep-targeted interventions.

Definitive RBD diagnosis requires clinical assessment with video-polysomnographic demonstration of RWA and dream-enactment^20–22^, combining multiple electrophysiological (EEG, EOG, EMG) and cardio-respiratory signals, with technician-assisted acquisition and expert interpretation to determine sleep stages and annotate events^21^. This workflow suits clinical diagnosis but is difficult to deploy at scale. Questionnaires offer inexpensive first-stage screening, but their performance is variable and depends on symptom awareness and a bedpartner. In early PD, sensitivity is limited, and their use has been discouraged^23^. Accordingly, the MDS research criteria for prodromal PD assign questionnaire-based RBD a positive likelihood ratio of only 2.8, versus about 130 for polysomnography-confirmed RBD^3,5^. In community screening, 33-43% of identified individuals did not have RBD on diagnostic video-polysomnography, despite combined questionnaires and clinical interview^8,24^. An objective home test need not replace polysomnography to address this bottleneck. It could provide second-stage objective screening after initial risk assessment (screening questionnaire, autonomic symptoms, genetic risk) and prioritise individuals for specialist assessment or support cohort stratification. This requires a complete pathway from self-application to an interpretable participant-level output, operating with minimal human intervention.

Deep-learning models such as U-Sleep can stage sleep from EEG and EOG inputs across heterogeneous polysomnography datasets in controls^25^ and in neurodegenerative diseases^26^. With ear-EEG electrodes, clinically useful sleep-stage information is retained in healthy controls^27,28^. REM sleep is difficult to identify in RBD, as the loss of atonia introduces muscle activity and movement artefacts, so epochs can resemble wakefulness^22^. Using standard polysomnography or reduced sensor sets, algorithms have quantified RWA from conventional chin and arm EMG placements and classified RBD^29–33^. Nevertheless, this automation is partial and begins only after skilled human-dependent acquisition or annotation. Reduced home systems still require separate sensors for sleep staging and RWA. Conventional submental (chin) EMG relies on electrodes that are susceptible to artefact from snoring, breathing, movement and electrode detachment, all of which must be identified or excluded during RWA quantification^22,32^. A system intended for RBD detection must accommodate fragmented and pathological sleep, enable high-quality EMG acquisition and allow self-application, while remaining tolerable for an older target population, especially as multiple nights may be needed for early diagnosis and follow-up.

We discovered that a single bipolar ExG derivation recorded from two electrodes behind the ears meets the electrophysiological requirements for RBD detection, capturing both the cortical activity needed for sleep staging and the muscle activity needed for RWA quantification, with optional EOG and arm-EMG channels adding information when required. We validated this finding and integrated the method into an end-to-end, fully automated system that uses mastoid ExG from self-applied recordings for automated sleep staging and RWA quantification, delivering RBD classification without manual annotations or review. We first trained a sleep-staging model for 1-channel ExG and fine-tuned it for sleep in people with Parkinson’s disease and isolated RBD using heterogeneous public and research polysomnographic datasets. We then utilised simultaneously acquired mastoid ExG and conventional polysomnography for further fine-tuning, and validated mastoid- and arm-derived RWA against blinded expert quantification from established chin and flexor digitorum superficialis EMG recordings. Finally, we assessed participant-level RBD classification and night-to-night repeatability in independent self-applied home recordings. By comparing mastoid-only, mastoid-EOG and full configurations with arm-EMG, we examined the trade-off between application burden, staging performance and diagnostic information. The resulting system is intended to support objective RBD screening and research stratification while providing a measurement basis for future longitudinal studies in synucleinopathies.

## Results

We developed a fully automated system that detects REM sleep behaviour disorder (RBD) from a self-applied pair of ExG electrodes without manual scoring or annotation at any stage. Mastoid signals (M1, M2), alone or with electrooculography (EOG1, EOG2), are sleep staged by a fine-tuned deep neural network, and high-confidence stage R sleep is passed for estimation of REM sleep without atonia (RWA) using bipolar mastoid (mastoid-EMG) alone or with bilateral flexor digitorum superficialis electromyography (FDS-EMG), before finally a classifier returns an RBD decision. In self-applied home recordings from 76 participants, the 1-channel system identified RBD with an area under the curve of 0.95 (sensitivity 94%, specificity 86%).

The system was developed and validated across four datasets. One- and 2-channel models were trained from scratch (Base Dataset), then fine-tuned to the target population (PACE-CBC Dataset), adapted to inferior electrode placements and home devices (Dual Dataset), and finally deployed on the Home Dataset of self-applied recordings. Demographic, clinical and sleep-staging metrics are presented in **Table 1**. Controls in PACE-CBC had high RBDSQ scores and apnoea-hypopnoea indices, as most were recruited from screening studies.

**Table 1.** Cohort characteristics across the three main datasets. Data are shown for the PACE-CBC, the Dual (concurrent PSG and home device) and Home Datasets, stratified by diagnostic group: controls (C), isolated REM sleep behaviour disorder (iRBD), Parkinson’s disease without RBD (PD^RBD-^) and Parkinson’s disease with RBD (PD^RBD+^). Not shown are small clinical subgroups of individuals with Multiple System Atrophy (n = 3 in PACE-CBC, 1 in Dual), Dementia with Lewy Bodies (n = 1 in PACE-CBC) and Pure Autonomic Failure (n = 8 in PACE-CBC, 3 in Dual, 1 in Home) and individuals with Parkinson’s disease in whom RBD status could not be established by polysomnography (n = 4 in PACE-CBC, 3 in Dual). In the Dual Dataset, demographic and manually scored variables are identical across the two signals and are reported once, while automated (model-derived) sleep metrics are reported on two lines per cell with the PSG signal (upper) and the home-recording signal (lower). The Dual Dataset contained no iRBD participants. Home recordings were not manually scored. Continuous variables are given as median [IQR] for demographic and clinical measures and as mean (SD) for sleep-architecture measures; categorical variables as n (%). AHI, apnoea-hypopnoea index; RBDSQ, REM Sleep Behaviour Disorder Screening Questionnaire; ESS, Epworth Sleepiness Scale; MoCA, Montreal Cognitive Assessment; TST, total sleep time.

| Characteristic | PACE-CBC Dataset |  |  |  | Dual Dataset |  |  | Home Dataset |  |  |  |
| --- | --- | --- | --- | --- | --- | --- | --- | --- | --- | --- | --- |
|  | Control | iRBD | PD<sup>RBD-</sup> | PD<sup>RBD+</sup> | Control | PD<sup>RBD-</sup> | PD<sup>RBD+</sup> | Control | iRBD | PD<sup>RBD-</sup> | PD<sup>RBD+</sup> |
| **Demographics & clinical** |  |  |  |  |  |  |  |  |  |  |  |
| N (nights) | 114 | 146 | 80 | 64 | 29 | 23 | 21 | 45 | 75 | 32 | 57 |
| N (unique subjects) | 114 | 146 | 59 | 64 | 29 | 21 | 21 | 19 | 27 | 15 | 21 |
| Age at recording (years) | 66.0<br>[57.9-71.6] | 67.1<br>[61.7-71.2] | 65.7<br>[61.2-70.7] | 69.3<br>[62.6-73.4] | 70.7<br>[65.2-76.0] | 67.0<br>[62.7-70.9] | 68.0<br>[63.4-73.5] | 66.5<br>[54.6-74.8] | 67.2<br>[63.4-69.8] | 69.2<br>[65.2-74.9] | 74.4<br>[68.3-77.7] |
| Sex (female, n (%)) | 33 (28.9%) | 23 (15.8%) | 22 (27.5%) | 22 (34.4%) | 11 (37.9%) | 8 (34.8%) | 10 (47.6%) | 23 (51.1%) | 12 (16.0%) | 5 (15.6%) | 20 (35.1%) |
| BMI (kg/m<sup>2</sup>) | 25.7<br>[23.6-27.8] | 25.2<br>[23.2-27.8] | 25.7<br>[23.1-28.4] | 24.6<br>[22.3-27.9] | 26.2<br>[23.7-28.7] | 25.8<br>[23.6-27.0] | 23.9<br>[22.9-27.2] | 23.5<br>[22.0-29.4] | 25.8<br>[23.7-27.5] | 25.4<br>[22.9-26.1] | 24.4<br>[22.9-25.2] |
| Apnoea-Hypopnea Index (AHI) | 10.4<br>[2.8-21.0] | 4.6<br>[0.8-12.8] | 9.3<br>[3.1-19.5] | 6.7<br>[2.8-13.2] | 17.8<br>[4.7-35.9] | 10.2<br>[4.9-26.9] | 10.3<br>[3.9-13.1] | - | 5.7<br>[0.2-11.6] | 10.2<br>[8.0-31.5] | 12.2<br>[3.7-15.8] |
| Epworth Sleepiness Scale (ESS) | 6.0<br>[4.0-9.0] | 6.0<br>[4.0-9.0] | 6.0<br>[4.0-9.0] | 6.0<br>[4.0-10.0] | 5.0<br>[4.0-7.0] | 6.0<br>[3.2-8.8] | 5.0<br>[3.0-8.0] | 5.0<br>[2.0-6.0] | 6.0<br>[2.0-9.0] | 5.0<br>[3.0-8.2] | 5.0<br>[3.0-9.2] |
| RBDSQ score | 7.0<br>[4.0-10.0] | 10.0<br>[8.8-11.0] | 3.0<br>[2.0-4.0] | 6.5<br>[4.0-9.0] | 3.0<br>[2.0-4.2] | 3.0<br>[2.0-4.0] | 6.0<br>[4.0-8.0] | 1.0<br>[1.0-2.5] | 10.0<br>[10.0-11.0] | 3.0<br>[1.0-4.0] | 8.0<br>[4.0-12.0] |
| Sniffin' Sticks 12-item score | - | 7.0<br>[5.0-8.0] | - | - | - | - | - | - | - | - | - |
| Sniffin' Sticks 16-item score | 11.0<br>[9.0-13.0] | 7.0<br>[6.0-10.0] | 7.0<br>[6.0-10.0] | 6.9<br>[5.2-8.9] | 11.0<br>[9.0-13.0] | 6.0<br>[5.0-9.5] | 7.0<br>[6.0-9.0] | 12.0<br>[11.0-13.0] | 6.0<br>[5.0-8.0] | 6.2<br>[5.0-10.0] | 6.0<br>[5.0-7.8] |
| MoCA total score | 27.5<br>[26.0-29.0] | 27.0<br>[26.0-29.0] | 28.0<br>[26.0-29.0] | 27.0<br>[26.0-28.0] | 27.0<br>[25.0-28.0] | 28.0<br>[26.8-29.0] | 27.0<br>[26.0-28.0] | 28.0<br>[27.5-29.0] | 28.0<br>[26.0-29.8] | 29.0<br>[27.0-30.0] | 27.0<br>[26.0-29.0] |
| **Sleep architecture – manual scoring** |  |  |  |  |  |  |  |  |  |  |  |
| Total sleep time (min) | 376.6 (76.5) | 394.6 (71.0) | 376.6 (75.1) | 392.0 (71.9) | 365.8 (87.2) | 369.0 (73.0) | 386.4 (87.2) | - | - | - | - |
| Sleep efficiency (%) | 79.7 (13.6) | 81.4 (12.6) | 77.2 (12.9) | 76.4 (14.2) | 73.5 (13.7) | 73.1 (12.9) | 72.9 (15.5) | - | - | - | - |
| Sleep onset latency (min) | 12.5 (14.3) | 30.4 (95.6) | 10.5 (11.3) | 19.7 (72.3) | 18.2 (18.4) | 13.1 (16.2) | 14.5 (14.9) | - | - | - | - |
| REM latency (min) | 123.1 (86.5) | 117.1 (80.3) | 143.7 (88.5) | 160.8 (100.1) | 156.4 (95.4) | 176.1 (85.5) | 204.5 (125.4) | - | - | - | - |
| N1 fraction of TST (%) | 21.7 (11.8) | 19.3 (9.3) | 13.5 (8.4) | 14.0 (9.2) | 23.4 (13.6) | 17.0 (8.7) | 18.9 (11.1) | - | - | - | - |
| N2 fraction of TST (%) | 49.6 (10.5) | 46.9 (9.2) | 54.5 (10.7) | 57.1 (8.4) | 51.1 (11.7) | 58.8 (10.9) | 60.3 (8.3) | - | - | - | - |
| N3 fraction of TST (%) | 13.8 (8.1) | 16.6 (7.0) | 15.3 (9.6) | 13.0 (8.0) | 9.7 (6.6) | 10.0 (8.3) | 8.4 (5.6) | - | - | - | - |
| REM fraction of TST (%) | 15.0 (7.7) | 17.1 (6.5) | 16.6 (7.9) | 15.9 (7.5) | 15.9 (10.8) | 14.2 (6.1) | 12.4 (6.5) | - | - | - | - |
| Wake after sleep onset (min) | 82.7 (65.7) | 65.0 (42.8) | 92.0 (60.5) | 100.9 (68.7) | 111.7 (70.1) | 114.8 (70.1) | 121.1 (87.4) | - | - | - | - |
| Stage change count | 162.1 (70.6) | 143.7 (48.0) | 144.7 (56.1) | 138.8 (47.0) | 202.8 (91.2) | 155.3 (63.3) | 172.0 (47.5) | - | - | - | - |
| **Sleep architecture – automated scoring** |  |  |  |  |  |  |  |  |  |  |  |
| Total sleep time (min) | 366.7 (75.9) | 372.8 (69.5) | 377.6 (69.7) | 383.1 (72.1) | 349.8 (74.8)<br>353.1 (76.6) | 334.8 (109.2)<br>335.9 (108.1) | 329.3 (114.1)<br>326.5 (113.4) | 402.9 (56.6) | 385.7 (68.2) | 331.2 (100.5) | 363.4 (71.6) |
| Sleep efficiency (%) | 77.9 (13.9) | 77.3 (13.0) | 77.3 (12.6) | 74.5 (13.9) | 74.0 (14.0)<br>75.0 (14.2) | 73.9 (13.3)<br>74.3 (12.3) | 72.8 (16.6)<br>72.4 (16.1) | 84.9 (9.1) | 78.7 (11.1) | 72.4 (19.4) | 74.9 (12.3) |
| Sleep onset latency (min) | 14.0 (16.5) | 32.9 (93.2) | 11.3 (11.3) | 19.8 (66.4) | 21.3 (24.0)<br>19.4 (20.1) | 11.6 (12.2)<br>11.0 (12.4) | 16.1 (22.1)<br>13.2 (18.2) | 21.3 (12.2) | 38.0 (29.8) | 35.2 (45.6) | 45.9 (43.9) |
| REM latency (min) | 129.3 (89.5) | 117.6 (78.9) | 137.3 (84.6) | 150.7 (76.6) | 165.4 (83.5)<br>173.2 (99.9) | 165.8 (90.3)<br>154.9 (81.0) | 139.4 (64.3)<br>144.4 (97.3) | 74.0 (38.8) | 94.0 (54.0) | 130.8 (87.3) | 136.2 (77.1) |
| N1 fraction of TST (%) | 16.1 (10.2) | 15.4 (8.0) | 12.7 (8.5) | 12.2 (9.3) | 17.8 (12.1)<br>19.5 (13.1) | 14.2 (9.1)<br>14.8 (8.7) | 11.8 (9.4)<br>13.8 (8.5) | 12.6 (5.5) | 13.3 (6.0) | 11.5 (4.5) | 10.7 (4.9) |
| N2 fraction of TST (%) | 56.7 (10.8) | 55.3 (10.2) | 58.3 (11.5) | 61.4 (10.6) | 58.2 (12.4)<br>55.6 (10.9) | 60.9 (10.0)<br>59.9 (9.9) | 61.7 (12.5)<br>61.2 (12.8) | 54.2 (8.0) | 55.9 (10.0) | 59.9 (12.4) | 59.5 (10.3) |
| N3 fraction of TST (%) | 10.3 (7.4) | 12.6 (7.6) | 12.6 (8.9) | 11.7 (8.5) | 8.3 (7.2)<br>9.4 (7.8) | 12.7 (10.2)<br>13.1 (9.9) | 12.3 (8.8)<br>12.6 (9.3) | 13.9 (5.9) | 14.2 (9.7) | 12.5 (7.9) | 11.6 (8.7) |
| REM fraction of TST (%) | 16.9 (7.8) | 16.7 (7.1) | 16.4 (8.8) | 14.7 (7.7) | 15.7 (10.6)<br>15.3 (10.2) | 12.2 (6.8)<br>12.2 (6.5) | 14.2 (7.9)<br>12.3 (7.8) | 19.2 (5.7) | 16.6 (6.7) | 16.1 (6.8) | 18.1 (8.4) |
| Wake after sleep onset (min) | 89.3 (65.1) | 82.6 (50.7) | 93.0 (63.1) | 107.4 (63.3) | 98.6 (61.7)<br>95.4 (63.6) | 97.5 (76.3)<br>97.1 (73.1) | 111.2 (88.8)<br>117.2 (88.3) | 49.9 (76.2) | 61.6 (49.1) | 76.5 (66.9) | 68.0 (49.2) |
| Stage change count | 139.5 (57.9) | 138.0 (44.4) | 127.1 (45.2) | 123.6 (41.0) | 155.0 (70.7)<br>167.8 (73.9) | 125.6 (65.2)<br>132.6 (73.1) | 117.3 (52.7)<br>125.0 (53.6) | 142.4 (45.4) | 147.6 (43.2) | 119.9 (42.6) | 120.5 (30.8) |

### Sleep staging models for minimal electrode montages

An ensemble across 2-channel combinations using the Base Model, reached a mean Cohen’s κ of 0.83 (median 0.85) against human scoring on the Base Dataset, with individual 2-channel combinations ranging from κ 0.801 to 0.825. Applied without adaptation to the clinical PACE-CBC Dataset (**Table 1, PACE-CBC Dataset**), agreement fell to κ 0.66 (stage R mean F1-score 0.87 to 0.76), reflecting domain shift from controls to an older population with neurodegenerative diseases. Fine-tuning the Base Model on the clinical dataset recovered most of this loss (**Figure 1a**), raising agreement to κ 0.73 (median 0.74) and stage R F1 to 0.84 (p = 6.44 × 10^-53^ and p = 2.80 × 10^-41^, respectively; two-sided Holm-adjusted Wilcoxon signed-rank test), approaching human inter-rater agreement (κ ≈ 0.76 in controls^34^ and ≈ 0.61 in PD^35^).

**Figure 1.**
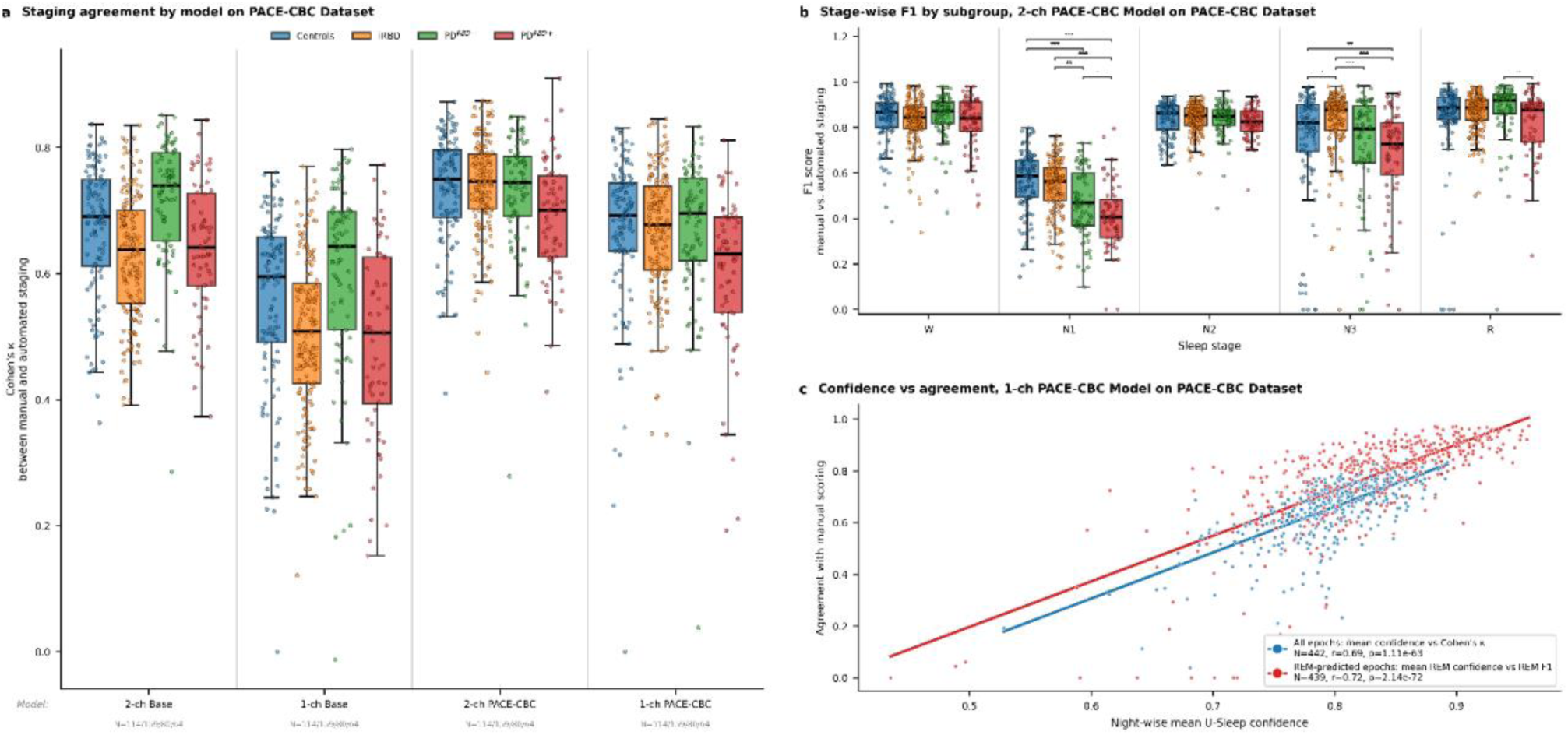
Sleep staging performance. **a**, Per-night agreement (Cohen’s κ) between automated and human sleep staging with points coloured by clinical subgroup. The 2-channel and 1-channel Base Models were trained on the Base Dataset (montages extracted from 6542 polysomnograms) and reached κ = 0.66 and κ = 0.53 when applied to the clinical PACE-CBC Dataset, respectively. Fine-tuning on the PACE-CBC Dataset increased agreement to κ = 0.73 for the 2-channel model, while the 1-channel model reached κ = 0.65 when tested on the PACE-CBC Dataset. **b**, Stage-wise, per-night F1 scores for the 2-channel PACE-CBC Model on the PACE-CBC Dataset, by clinical subgroup. Staging remained accurate for stage R in patients with RBD, with N1 as the most difficult stage across all groups. Significance indicated by stars (* < 0.05, ** < 0.01, *** < 0.001). **c**, Association between per-night confidence and κ (red) and between epoch-wise stage R confidence and stage R F1 (blue) for the 1-channel fine-tuned model. Higher model confidence corresponded to closer agreement with human scoring (r = 0.69 and r = 0.72, respectively).

Concurrent in-laboratory polysomnography and home device recordings were acquired (84 nights), with home device electrodes applied secondarily, but as close to the intended positions as possible (**Table 1, Dual Dataset**). Applying the 2-channel PACE-CBC Model directly to the home device signals yielded a mean κ of 0.62 against human scoring of the polysomnography signals. Further fine-tuning on the home device recordings (Dual Model) raised the mean κ to 0.67. Automated scoring of the polysomnography and home device signals, both using the Dual Model, agreed closely (κ = 0.83), and the remaining gap may be explained by differences in electrode placements and device properties. For the 1-channel PACE-CBC Model on the Dual Dataset, mean κ was 0.50 prior to fine-tuning, recovering to a mean κ of 0.54.

Clinical sleep metrics, including total sleep time (TST), sleep efficiency (SE) and wake after sleep onset (WASO), were not significantly different from human scoring in PACE-CBC when using the 1-channel or 2-channel model (p > 0.165 for all). The 2-channel model was numerically closer to human scoring, and both models were significantly different in sleep stage changes and some other sleep metrics.

Using the 1-channel PACE-CBC model on the PACE-CBC Dataset, the U-Sleep confidence measure correlated with per-night κ (r = 0.69, p = 1.11 × 10^-63^) and, for stage R specifically, with stage R F1 (r = 0.72, p = 2.14 × 10^-72^, **Figure 1c**). The prespecified 80% stage R confidence threshold for selecting high-confidence epochs for RWA estimation discarded, on average, 34.8% of stage R epochs per night while raising stage R precision from 78.3% to 87.3%, with 6 recordings (1.3%) having no high-confidence REM (**Table 2**).

**Table 2.**
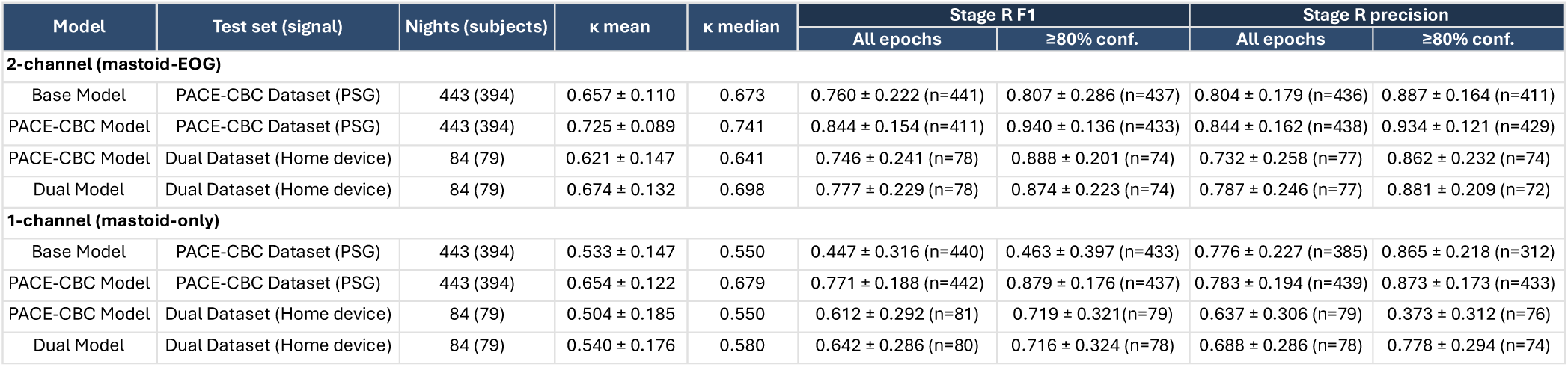
Sleep-staging agreement for the 1-channel and 2-channel models. The *Base Model* is trained on the multi-site Base Dataset consisting of controls, while the *PACE-CBC Model* is the Base Model fine-tuned on the clinical PACE-CBC dataset, and the *Dual Model* is the PACE-CBC Model further fine-tuned on the Dual Dataset. Dual Dataset rows apply each model to the signals acquired with the home device, scored against human annotations from the concurrent polysomnography. ‘All epochs’ includes every stage R epoch while ‘≥80% conf.’ restricts to stage R epochs where model confidence is at least 0.80. Stage R precision is undefined for a night in which the model predicted no stage R epochs and is excluded from that cell’s mean. In contrast, F1 is undefined only when REM is absent from both human scorings and model predictions and evaluates to 0 when either human or model identifies REM while the other does not, resulting in differing sample sizes (n) between these metrics.

### Estimates of REM sleep without atonia using mastoid-EMG and arm-EMG

Across PACE-CBC, automated RWA estimates correlated strongly with manual SINBAR scoring. Automated mastoid-EMG RWA correlated strongly with human SINBAR chin-EMG scoring on a per-night basis (r = 0.82, p = 2.88 × 10^-34^, n = 136; **Figure 2b**), and bilateral FDS-EMG RWA correlated strongly with the SINBAR phasic FDS-EMG score (r = 0.93, p = 8.69 × 10⁻^31^, n = 69; **Figure 2c)**.

**Figure 2.**
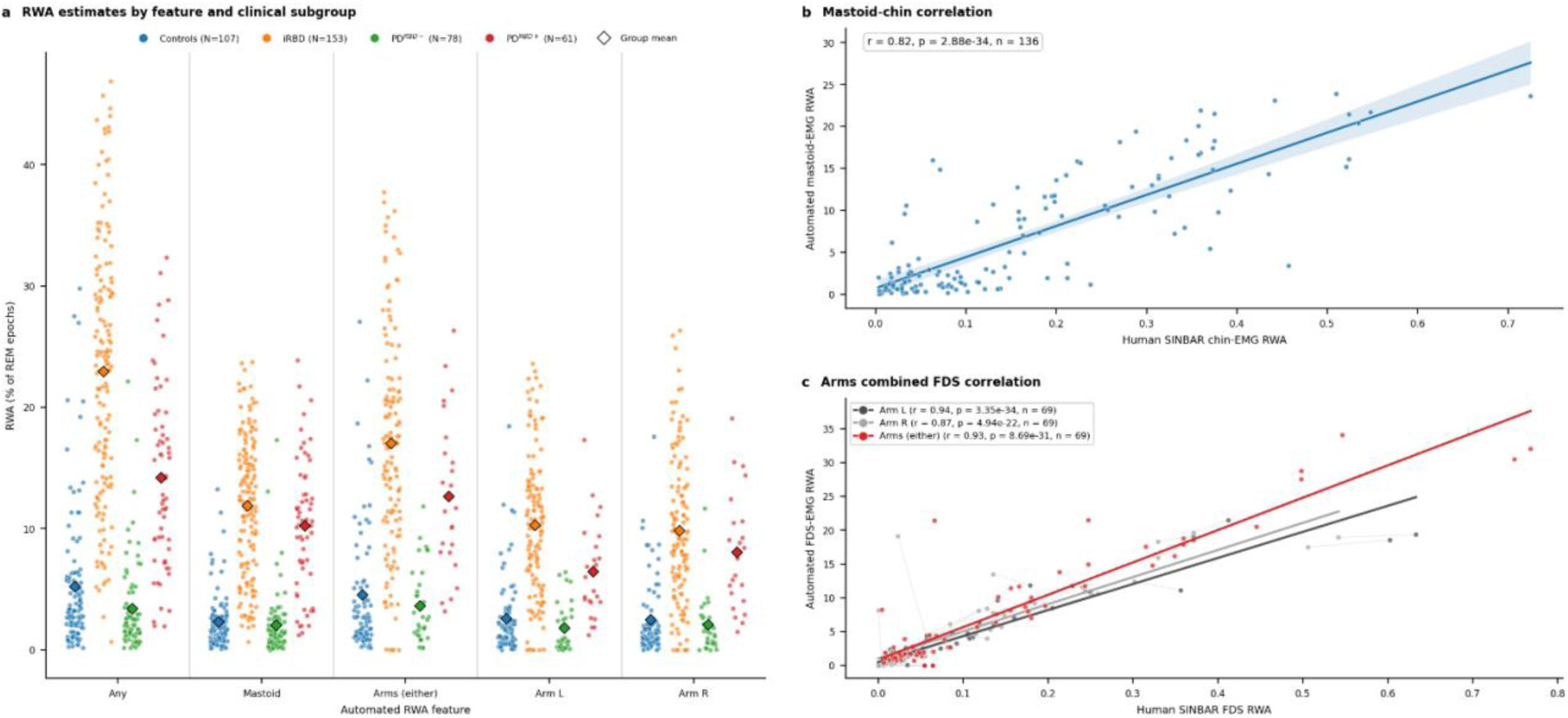
REM sleep without atonia (RWA) estimates from mastoid-EMG and arm-EMG. **a**, Automated RWA estimates in high-confidence (≥80%) stage R from the PACE-CBC Dataset, by feature and clinical subgroup (Controls, iRBD, PD^RBD-^, PD^RBD+^). Features reflect estimated RWA across three channels (mastoid, left arm FDS, right arm FDS), presented as *any* (combined activity), *mastoid* (bipolar M1-M2), *arms (either)* (bilateral combined), *arm L* (left arm), and *arm R* (right arm); diamonds indicate group means. **b**, Per-night correlation between automated mastoid-EMG RWA and human SINBAR scoring of chin-EMG RWA (r = 0.82, p = 2.88 × 10^-34^, n = 136). **c**, Per-night correlation between automated FDS-EMG RWA and human SINBAR scoring of FDS-EMG, shown for the bilateral *arms (either)* estimate (red: r = 0.93, p<0.001, n = 69) and for the left and right arm separately (grey: r = 0.94 and 0.87), with each subject’s left and right values joined by a thin line. Panels b and c show ordinary least-squares fit lines with a 95% confidence band in panel b. Automated estimates fall below the human scores at roughly half the SINBAR magnitude. That the mastoid–chin correlation remains strong indicates that the crosshead mastoid derivation picks up residual chin activity, or that the degree of RWA in the mastoid region is physiologically correlated with that in the chin.

RWA differed markedly across clinical subgroups (**Figure 2a**). The iRBD and PD^RBD+^ showed substantially higher RWA than those without RBD (p < 0.05), and among RBD groups, iRBD showed numerically higher RWA than PD^RBD+^ across all features (p > 0.05, Holm-corrected).

### Diagnostic performance on self-applied home recordings and in-lab PSG

The complete pipeline classified RBD directly from self-applied home recordings, with no manual annotation at any stage. Using the 1-channel mastoid-only montage, RBD was detected at subject-level with an AUC of 0.95 (sensitivity 94%, specificity 86%; n = 76, 48 RBD / 28 non-RBD; **Figure 3b**, **Table 3**). Adding EOG for staging (2-channel model) gave an AUC of 0.95, and additionally including arm-EMG gave 0.97.

**Figure 3.**
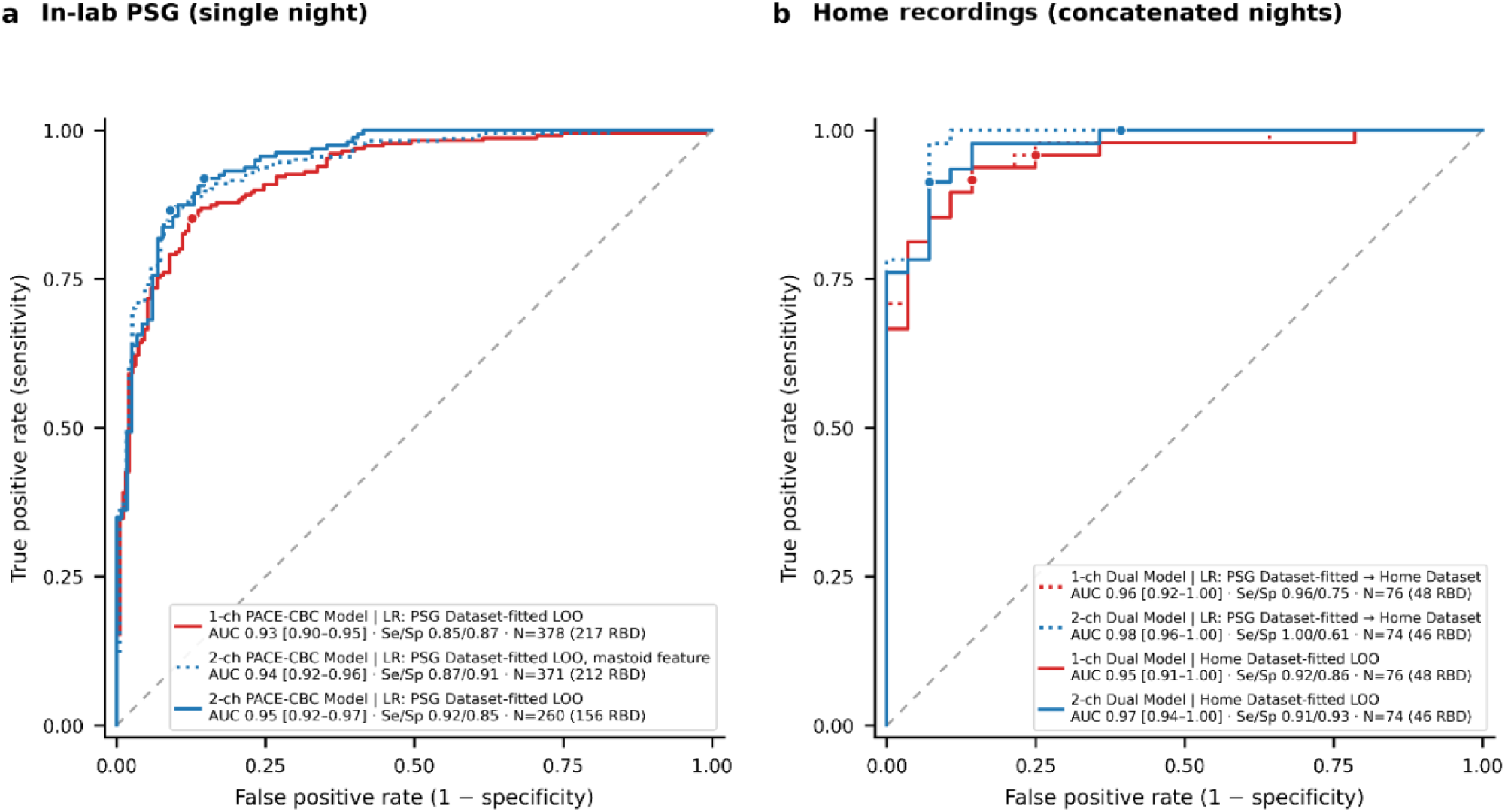
Diagnostic performance for detecting RBD in-lab and at home. Performance of the minimal 1-channel mastoid-only, 2-channel mastoid-EOG using mastoid RWA, and the full 2-channel mastoid-EOG with arm-EMG (combined mastoid and FDS RWA). **a**, Receiver-operating-characteristic (ROC) curves for classifying RBD versus non-RBD on single-night in-lab polysomnography (PACE-CBC Dataset), achieving AUC 0.93 (1-channel), 0.94 (2-channel, mastoid-only) and 0.95 (2-channel, full montage). **b**, ROC curves for self-applied home recordings using concatenated stage R across 1-3 nights, achieving AUC 0.95 (1-channel) and 0.97 (2-channel, full montage) when thresholds were fitted using leave-one-out regression. Filled circles mark the operating point for the reported sensitivity and specificity. The dashed diagonal indicates chance. All models used an 80% stage R confidence threshold.

**Table 3.** Diagnostic performance for classifying REM sleep behaviour disorder (RBD) from REM sleep without atonia. Performance of the 1-channel mastoid-only montage, the 2-channel mastoid-EOG montage using mastoid-RWA, and the 2-channel mastoid-EOG montage using mastoid-RWA and arm-RWA. Within each montage, ‘In-lab PSG (LOO)’ is leave-one-out cross-validation of the logistic regression across the in-lab PSG nights, staged with the PACE-CBC Model. ‘Home Dataset (PSG-transfer)’ applies the coefficients and the feature standardisation fitted on the in-lab PSG, unchanged, to the concatenated home recording nights staged with the Dual Model, with only the operating cut-off set on the home data. ‘Home Dataset (LOO)’ is leave-one-out within the home cohort. In-lab (PSG) sensitivity, specificity and confusion counts (TP / TN / FP / FN) are computed per analysed recording, with a subset of participants contributing more than one recording. All montages used the 80% stage R confidence threshold.

| Evaluation | Staging model | N (RBD/non-RBD) | AUC [95% CI] | Sensitivity [95% CI] | Specificity [95% CI] | Balanced accuracy | TP/TN/FP/FN |
| --- | --- | --- | --- | --- | --- | --- | --- |
| **1-channel PACE-CBC Model** |  |  |  |  |  |  |  |
| In-lab PSG (LOO) | PACE-CBC Model | 378 (217/171); 420 rec. | 0.93 [0.90-0.95] | 0.86 [0.81-0.90] | 0.87 [0.82-0.91] | 0.87 | 197/166/24/33 |
| Home Dataset (PSG-transfer) | Dual Model | 76 (48/28) | 0.96 [0.92-1.00] | 0.94 [0.83-0.98] | 0.86 [0.69-0.94] | 0.90 | 45/24/4/3 |
| **Home Dataset (LOO)** | **Dual Model** | **76 (48/28)** | **0.95 [0.91-1.00]** | **0.94 [0.83-0.98]** | **0.86 [0.69-0.94]** | **0.90** | **45/24/4/3** |
| **2-channel PACE-CBC with/without arms** |  |  |  |  |  |  |  |
| *Mastoid RWA (without arms)* |  |  |  |  |  |  |  |
| In-lab PSG (LOO) | PACE-CBC Model | 371 (212/170); 413 rec. | 0.94 [0.92-0.96] | 0.87 [0.82-0.91] | 0.91 [0.86-0.94] | 0.89 | 195/172/17/29 |
| Home Dataset (PSG-transfer) | Dual Model | 76 (47/29) | 0.96 [0.91-1.00] | 0.94 [0.83-0.98] | 0.90 [0.74-0.96] | 0.92 | 44/26/3/3 |
| Home Dataset (LOO) | Dual Model | 76 (47/29) | 0.95 [0.90-1.00] | 0.94 [0.83-0.98] | 0.90 [0.74-0.96] | 0.92 | 44/26/3/3 |
| *Mastoid + arm RWA (fixed, with arms)* |  |  |  |  |  |  |  |
| In-lab PSG (LOO) | PACE-CBC Model | 260 (156/107); 276 rec. | 0.95 [0.92-0.97] | 0.92 [0.87-0.95] | 0.85 [0.78-0.91] | 0.89 | 147/99/17/13 |
| Home Dataset (PSG-transfer) | Dual Model | 74 (46/28) | 0.98 [0.96-1.00] | 0.98 [0.89-1.00] | 0.93 [0.77-0.98] | 0.95 | 45/26/2/1 |
| Home Dataset (LOO) | Dual Model | 74 (46/28) | 0.97 [0.94-1.00] | 0.91 [0.80-0.97] | 0.93 [0.77-0.98] | 0.92 | 42/26/2/4 |

Applied to single-night in-lab polysomnography (PACE-CBC Dataset), the 1-channel montage detected RBD with an AUC of 0.93, slightly lower than the 2-channel montage including arm-EMG, which achieved 0.95. At home, the benefit from added channels was minimal with AUCs of 0.95, 0.95 and 0.97 for 1-channel, 2-channel and 2-channel-with-arms montages. Transferring the PACE-CBC-trained classifier to the Home Dataset reproduced home performance (AUC 0.96, 1-channel).

### Multiple nights provide insights into intraindividual variability

Across participants with three home nights, high-confidence REM was equally available on every night after cleaning (91.9%, 93.5% and 88.7%, p = 0.459), and among scorable nights no RWA estimate varied with night order (lowest FDR-corrected p = 0.290), indicating no first-night effect in RWA. Single-night accuracy of the 1-channel model was numerically higher on the second night than the first (AUC 0.866, 0.951 and 0.905 for nights 1 to 3) but did not survive correction (uncorrected p = 0.045, Holm-corrected 0.136).

Mastoid-RWA showed good test-retest reliability in the pooled population, with a single-night ICC of 0.77 (95% CI 0.62 to 0.86), a standard error of measurement of 3.0 percentage points and a smallest detectable change of 8.2 percentage points (**Figure 4a**). Reliability differed markedly between groups. It was higher in the two groups without RBD (ICC 0.72 in Controls and 0.95 in PD^RBD−^) than in the two RBD groups (ICC 0.19 in iRBD and 0.63 in PD^RBD+^), consistent with larger night-to-night fluctuation of RWA where values are high (**Figure 4b**).

**Figure 4.**
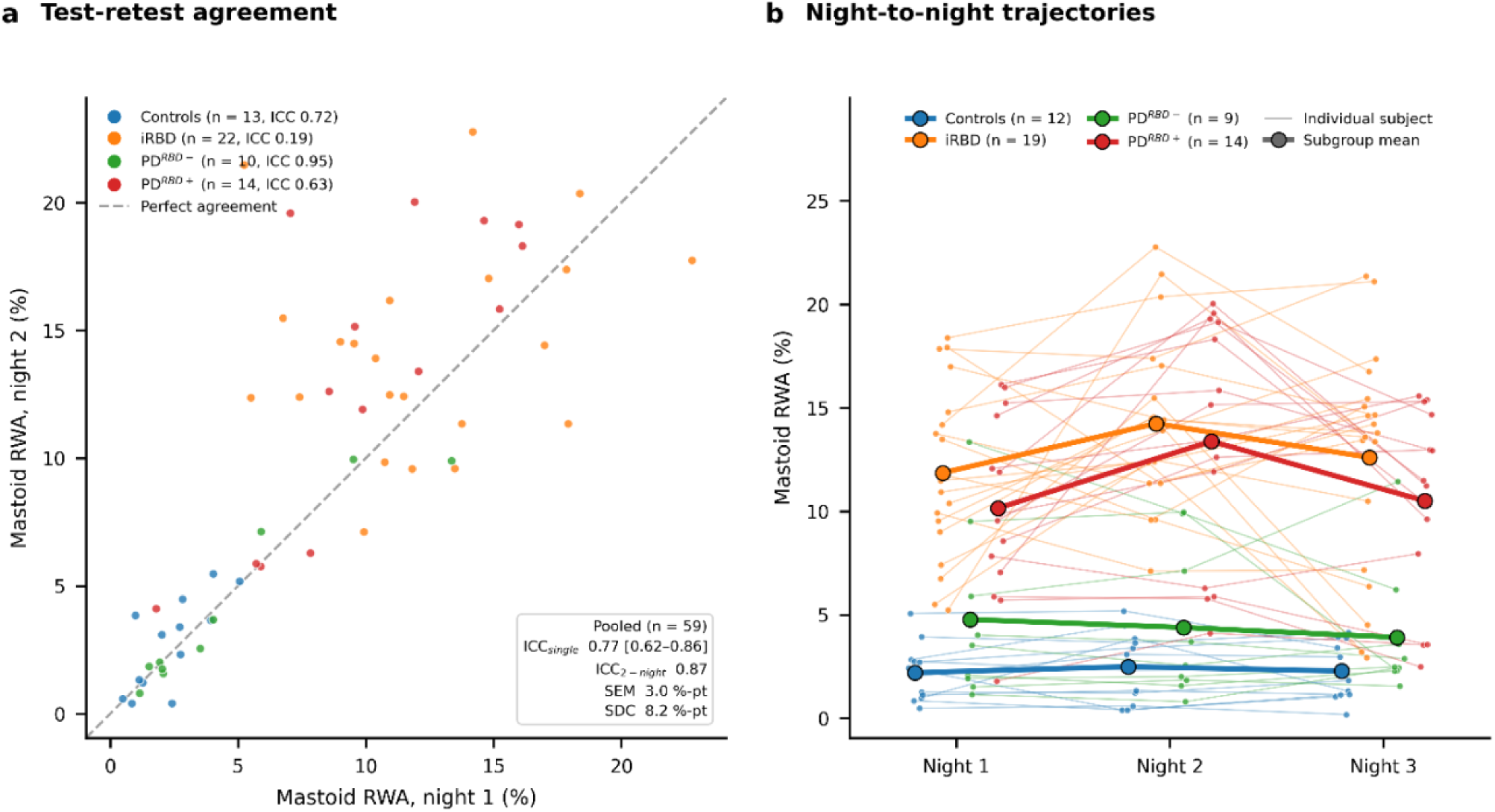
Variability of automated mastoid REM sleep without atonia (RWA) across home nights. Mastoid RWA was estimated from high-confidence stage R in self-applied home recordings and grouped by diagnosis (Controls, iRBD, PD^RBD−^, PD^RBD+^). **a**, Agreement between night 1 and night 2 (pooled n = 59), with each point representing one participant and the dashed line indicating perfect agreement. Across the full range of RWA, single-night reliability was good (pooled ICC 0.77, 95% CI 0.62 to 0.86) with a standard error of measurement of 3.0 percentage points and a smallest detectable change of 8.2 percentage points for the pooled group. Within diagnostic groups, single-night reliability was high in participants without RBD (ICC 0.72 in Controls and 0.95 in PD^RBD−^) and lower in the RBD phenotypes (ICC 0.19 in iRBD and 0.63 in PD^RBD+^), consistent with greater night-to-night fluctuation of RWA where values are high. **b**, Night-to-night trajectories across the three home nights for each participant (thin lines) with subgroup means (bold), coloured by diagnosis. Because the low-RWA (no RBD) and high-RWA (RBD) ranges stay well separated across nights, a single night is sufficient to distinguish RBD from non-RBD, whereas the greater within-patient fluctuation of RWA in the RBD groups indicates that repeated nights may improve estimation of RWA burden for disease monitoring.

## Discussion

We demonstrate that a self-applied pair of behind-the-ear (mastoid) electrodes enables fully automated RBD detection through sleep staging and quantitative REM sleep without atonia (RWA) in a novel mastoid-EMG derivation, without manual scoring or assistance at any stage. To our knowledge, it is the first system to derive both automated staging and quantitative RWA from self-applied electrodes, achieving an AUC of 0.95 across 76 participants. This differs from previous simplified or modified polysomnography setups^29–31,33^ in the signal types and the degree of workflow automation. Previous methods all acquire chin-EMG with technician aid for montage and/or scoring and remain semi-automatic. While automating individual steps reduces workload, only full automation provides scalability and removes inter-rater variability, and the need for experts and technicians for every participant.

Reliable sleep staging is a prerequisite for home RWA analysis. The Base Model trained only on controls performed substantially worse in PACE-CBC, whereas fine-tuning the 2-channel model recovered performance to around the expected manual inter-rater ceiling, which is itself lower in PD than in controls. A single, consistently applied model removes between-rater variability across datasets, and per-epoch confidence provides a tool for selecting high-confidence R-sleep or rejecting unreliable nights. Adding electrooculography and ensemble staging improved sleep metric accuracy but not binary RBD classification. While 1-channel mastoid is sufficient for a screening decision, the richer montage is preferable for applications where accurate sleep architecture is needed.

The mastoid ExG channel acquires high-quality EEG, EMG and possibly EOG from accessible, off-the-shelf electrodes in a robust, low-burden montage without individualisation. Mastoid-RWA correlated strongly with manually scored chin-RWA, despite different anatomical sites. Its mechanism remains unresolved and is not previously described, possibly reflecting volume-conducted submental activity or co-activation across cranial muscles. Part of the ceiling on agreement is set by SINBAR itself, whose manual scoring is labour-intensive and rater-dependent, so consistency, diagnostic separation and prognostication, rather than one-to-one agreement, are the relevant benchmarks. The 1-channel model separated RBD with high accuracy, and although RWA detection is not equivalent to RBD diagnosis, our automated agreement with clinical RBD is high and matches or exceeds prior reports of RWA-RBD concordance^36^. As expected from different epoch rescoring (confidence vs manual) and temporal summation (continuous vs mini-epoch), our estimate was consistently about half of the manual value.

Beyond binary classification, RWA is increasingly viewed as a graded marker of neurodegeneration, with higher RWA predicting phenoconversion in iRBD^19,37^. Continuous quantification outperforms conventional epoch-based quantification for phenoconversion^18^, which discards information when RWA is reduced to 3-s binary events. Our continuous estimate revealed roughly twofold larger night-to-night fluctuations in established RBD than in controls. This adds to previous two-night in-lab studies in PD^38^ and defines how the measure should be used. A single night sufficed for a binary decision, while the physiological variability of RWA in RBD groups underlines that RWA quantification, especially in RBD, benefits from multiple nights, preferably at home, further sharpening its value as a biomarker for prognostication and progression tracking.

The continuity of RWA is illustrated by misclassified individuals. Of seven subjects misclassified by the 1-channel Home Model, two of the four false positives were PD^RBD-^ patients who showed RWA and REM behavioural events on recent polysomnography, albeit not fulfilling diagnostic criteria. These signs of emerging RBD were not present at baseline polysomnography six years earlier. Among three false negatives, one had converted from PD^RBD-^ to PD^RBD+^ within the preceding three years, and both iRBD patients showed no neurodegeneration on dopaminergic PET or cardiac sympathetic imaging at their baseline visit, one of whom remained free of neurodegeneration at 3- and 6-year follow-up, prompting re-evaluation. Some errors are therefore more compatible with early or ambiguous biology than with model failure, and the full 2-channel model with arm-EMG reduced the number of misclassifications to six.

The setting of a study matters. Our datasets include PD patients and controls with RBD symptoms identified through community screening. Community iRBD and PD^RBD+^ patients’ RBD is likely earlier, subtler and more variable, whereas retrospective validation in sleep clinics likely reflects a highly selected cohort of severe iRBD. Some misclassifications may reflect confounders, but the algorithms were deliberately kept naive to demographics, breathing, arousals, and movement artefact, keeping them fully automatic.

Home testing has reshaped sleep apnoea detection, cut waiting times and reduced per-patient costs^39^. In community screening for RBD, a reliable objective tool could reduce reliance on expert triage now used to filter referrals before polysomnography. It is itself imperfect, as only about 63% (78/124) of those referred for video-polysomnography during screening were confirmed to have RBD and true cases were missed^8,24^. Many community presentations mimic RBD, including untreated sleep apnoea, NREM parasomnias, PTSD-related nocturnal behaviours, and antidepressant use. Any screening method should be validated not only in diagnosed sleep-clinic iRBD patients and asymptomatic controls, where questionnaires already perform well, but in the screening setting where all participants have symptoms, yet only some have true RBD. Not all mimics are sufficiently represented in our dataset, although most controls came from community screening studies. Triage before polysomnography is needed, and across the wide span in precision and workload from a questionnaire-based "probable RBD" to gold-standard video-polysomnography^3,5^, a fully automated objective test could increase precision and reduce known biases, such as the sex bias of enactment-based instruments that under-detect women^40^. RWA tools also help stratify disease, defining the body-first subtype^15^ and the diffuse-malignant phenotype^13^, where RBD carried prognostic and post hoc treatment signals in PASADENA^16^. Since neuroprotective trials depend on large, well-characterised iRBD cohorts that are currently difficult to assemble^41^, scalable screening would speed their development.

Multimodal screening may provide synergistic effects. Single-channel mastoid captures the core electrophysiological features of the disease, while movement-based tools infer RBD indirectly from wrist actigraphy, lumbar accelerometry or nearables such as video or radar. Receiving no electrophysiological signals, they are generally agnostic to sleep stages and RWA. High-confidence REM staging could be transferred to movement-based screening tools that lack reliable staging, or RWA estimates could complement alpha-synuclein seed-amplification assays, which confirm pathology as a binary readout, but not neurodegeneration or graded prognosis. Such combinations may outperform diagnostic labels alone, particularly for diagnostic mimics and isolated RWA preceding overt RBD and align with emerging frameworks for a biological classification of PD pairing molecular detection of pathology with functional measures of neurodegeneration^42,43^.

Several limitations apply. The system has not been deployed as a true first-line screening tool, as PACE-CBC controls were pre-selected via expert interview and the Home Dataset was not acquired in community screening. The RWA algorithm and classifier were left naive to confounders requiring human input, and excluding possibly affected subjects would bias the cohort. The RWA estimates were not designed to reproduce SINBAR, so their calibration is a visualisation rather than a replication. Generalisation across hardware was not examined in detail, though several devices were used without device-specific preprocessing, and unlike the heterogeneous Base Dataset, the fine-tuning and validation datasets were predominantly European and male, reflecting the epidemiology of PD and iRBD. Unsupervised home acquisition introduces uncertainty about, for instance, electrode placement, which we did not survey, and produced failed or low-quality nights of uncertain cause. Finally, scalable prodromal screening raises the question of disclosing neurodegenerative risk before disease-modifying therapy exists, though prodromal individuals generally support early detection^44^.

By screening with accessible, self-applied and fully automated methods, objective RBD detection becomes deployable at scale, while continuous RWA estimates provide quantitative substrates for stratification and longitudinal follow-up. Single-channel mastoid ExG is a convenient and robust acquisition method for electrophysiology-based early detection, and if confirmed prospectively across centres in community screening, it could help build and monitor the large prodromal cohorts that neuroprotective trials now require, alongside existing screening methods and confirmatory polysomnography. As disease-modifying trials mature, the bottleneck shifts from recruitment to finding and monitoring the right people early.

## Methods

### Study design and datasets

Model development used retrospectively and prospectively collected in-laboratory video-polysomnography and concurrent home device recordings, and the primary validation used prospectively collected self-applied home recordings. The index test was the automated pipeline, and the reference standard was an expert diagnosis of REM sleep behaviour disorder established from diagnostic video-polysomnography as described below.

Four different datasets were obtained for the development, fine-tuning and validation of sleep staging models. For the Base Dataset, a publicly available dataset of almost 20,000 polysomnograms was searched for recordings with availability of bilateral mastoid (M1, M2) and eye electrodes (EOG1, EOG2), resulting in the identification of 6542 eligible polysomnograms from adult subjects with and without comorbid sleep disorders, all with manually annotated sleep stages according to either RCK or AASM guidelines. All datasets used were handled in accordance with their data use policies.

The *Base Model* for sleep-staging using mastoid and EOG was trained from scratch using an openly available multi-site dataset (*Base Dataset*, 6055 participants, 6542 nights) by extracting polysomnography channels. Three large research datasets acquired were then used for fine-tuning and validation. A polysomnography dataset at PACE and CBC (*PACE-CBC Dataset*, 394 participants, 443 nights) was used to fine-tune the *PACE-CBC Model* to the target population of patients with isolated REM sleep behaviour disorder (iRBD) and Parkinson’s disease (PD) and included subjects with iRBD (n = 146) and PD with or without RBD (n = 64 and 59, respectively). A subset of PACE recordings within the PACE-CBC contained concurrent in-laboratory polysomnography and technician-applied home-device recordings (*Dual Dataset*, 79 participants, 84 nights), allowing the *Dual Model* to be fine-tuned and the two signal types to be compared within the same night. Finally, a *Home Dataset* of 82 participants at PACE contributing 211 self-applied home-recording nights (1–3 per participant) constituted the clinical deployment setting in which the complete pipeline was evaluated.

For the fine-tuned PACE-CBC Models, a total of 443 polysomnograms from two research centres were collected. The PACE recordings were collected at the Lundbeck Foundation Parkinson’s Disease Research Center at Aarhus University Hospital (PACE), and data were collected retrospectively and prospectively from ongoing longitudinal studies including patients with iRBD, PD and controls^15,45,46^. Patients with iRBD are partly from community screening and local sleep clinics. The Cologne-Bonn recordings were collected at the University of Cologne and University of Bonn (CBC) and consisted of retrospectively collected data from iRBD, PD and controls^24,47^, with iRBD cases identified mostly through community screening. For both datasets, most controls were initially included in previously published studies of community-based screening for iRBD or ongoing studies of early and prodromal PD. Controls from screening were identified based on questionnaires, telephone interviews and expert opinion, where subjects deemed to have possible iRBD underwent diagnostic video-polysomnography. If the suspicion of iRBD was ruled out based on video-polysomnography, the subject was included in the datasets and labelled as control.

The dual-device dataset (Dual Dataset) and the home-based dataset (Home Dataset) were collected prospectively at PACE. The dual-device dataset was used to fine-tune the Dual Model and comprised nights where diagnostic video-polysomnography and home device recording were recorded simultaneously (84 nights, 79 subjects). All polysomnographies in the Dual Dataset were also included in the PACE-CBC Dataset using the same subject splits. On concurrent recordings, the home device electrodes were placed inferiorly to the polysomnography electrodes, typically with mastoid electrodes placed slightly lower, and EOG electrodes placed slightly lateral to the standard AASM placement. During dual-device montage, all electrodes were mounted by technicians. This fine-tuning step was carried out to increase robustness towards slightly misplaced electrodes and device differences.

The Home Dataset was collected at PACE between 2023 and 2026. Equipment and written instructions were sent directly to the participants or handed out during research visits. Participants were allowed to get assistance from relatives in handling the electrodes and device, but no assistance from technicians was provided apart from providing information prior to the home recordings. Participants performed 1-3 nights of data acquisition depending on the study and the participant’s availability.

### Inclusion and exclusion criteria

Participants in the PACE-CBC Dataset were included from previous and ongoing studies applying slightly different eligibility rules. All participants were adults aged between 40 and 85, and excluded if they had epilepsy with nocturnal seizures, narcolepsy, major comorbidity (heart, kidney or respiratory failure, active cancer treatment), major stroke, alcohol abuse and major psychiatric conditions (except depression). Controls were defined as participants without any of the studied conditions (iRBD, PD or dementia) and a video-polysomnography demonstrating no evidence of RBD or both PE2I-PET and MIBG-SPECT imaging demonstrating no signs of neurodegeneration. Controls were not excluded based on symptoms of RBD or PD.

RBD was diagnosed using the ICSD-3 diagnostic criteria and an eligible video-polysomnography reviewed by a somnologist who was blinded to the outcome of the automated analysis^20^. In participants undergoing home-based measurements, the RBD diagnosis used as reference was based on a diagnostic video-polysomnography performed within 1 year from the home-based measurement. Antidepressant use was allowed if not inducing secondary RBD across all subgroups. RBD was found to be clearly induced by antidepressant use or PTSD without any signs of neurodegeneration, leading to exclusion from the evaluation of diagnostic performance.

PD was diagnosed according to the clinical Movement Disorder Society (MDS) criteria^48^. In participants with normal dopamine scans, the diagnosis of PD was dropped according to the MDS criteria, and these were considered as controls in the datasets. For cases with PD in whom RBD status could not be firmly established by polysomnography after review by two independent raters, the polysomnogram was kept for training of sleep staging models but was not applicable in the evaluations of diagnostic performance.

### Clinical assessments

Clinical assessment was carried out for research participants at PACE and CBC, including assessment of motor symptoms (MDS-UPDRS Part III), cognitive function (Montreal Cognitive Assessment, MoCA) and olfactory function (16- or 12-item Sniffin’ Sticks odour identification). Additionally, questionnaires concerning sleep symptoms were completed (REM behaviour disorder Screening Questionnaire and Epworth Sleepiness Scale). Clinical history was obtained, including the onset of dream enactment and/or motor symptoms.

### Ethics

The studies were approved by the Central Denmark Region Committees on Health Research Ethics (1-10-72-160-16) and by the ethics committee of Cologne University Hospital (19-1408). Prespecified analyses were not formally registered. Subjects provided informed, written consent according to the Declaration of Helsinki.

### Home devices and electrode montages

Home device recordings were performed using self-adhesive silver/silver chloride electrodes for measuring EEG, EOG and EMG signals. Electrodes were placed behind the ear at the left (M1) and right (M2) mastoid bone, below the left eye (EOG1) and above the right eye (EOG2). Two electrodes were placed on each arm 3-5 cm apart at the flexor digitorum superficialis, and a ground electrode was placed on the right side of the chest area, below the collarbone. Standard skin preparation was performed prior to electrode placement. Signal recording was performed using an amplifier platform, either the Mobita EEG amplifier (TMSi, Netherlands) or a similar in-house-built device. The device was placed in a pouch mounted on an elastic chest band. All recordings were collected at a sample rate of 1000 Hz and subsequently downsampled for analysis. No manual annotation was performed on the home device recordings.

### Video-polysomnography

In both research datasets (PACE, CBC), all video-polysomnographies were performed and manually annotated according to the AASM manual^21^. In brief, video-polysomnography was acquired using Somnomedics Somnoscreen Plus (Somnomedics, Germany) and included the standard 10-electrode EEG montage (10/20 system: F3, F4, C3, C4, O1, O2, M1, M2, Fpz as grounding, Cz as reference) with electrooculography (EOG1, EOG2). Electromyography (EMG) was recorded from the chin (Chin1, Chin2) and bilaterally from the anterior tibial muscles and the flexor digitorum superficialis muscles (FDS). Additionally, the recordings included electrocardiography, nasal pressure and flow monitoring, thoracic and abdominal respiratory effort, finger pulse oximetry, and synchronised audiovisual recording using infrared video. Manual PSG scoring was performed according to the AASM manual with sleep staging on 30-second epochs and annotation of arousals and respiratory events. Lights off and lights on were defined manually.

### Manual RWA quantification

Manual quantification of REM without atonia (RWA) was performed according to the SINBAR methodology on chin-EMG and FDS-EMG^22^. Recordings were discarded for manual quantification if the signal quality of the EMG channels was corrupted by technical artefacts or for chin-EMG by snoring during all available REM sleep. Recordings were also discarded if no REM sleep periods were present or all REM periods were severely affected by respiratory arousals. Prior to annotation of RWA, the REM sleep periods were rescored according to the SINBAR recommendations in 3-second mini-epochs with each period starting with a rapid eye movement and ending with either an arousal, N2 hallmark (sleep spindle or K-complex) or three consecutive minutes of REM sleep without rapid eye movements. Quantification of RWA was carried out blinded to the automated outputs of the algorithms.

### Data processing, cleaning, and rejection

To standardise the recordings and suppress artefacts, we applied a preprocessing pipeline organised into four phases. The recordings were preprocessed separately for the 1-channel and 2-channel models to ensure that information from the EOG electrodes did not affect the 1-channel (mastoid-only) model during cleaning. Throughout, rejected samples are not deleted but marked as missing (set to NaN), preserving the original timeline. For the 2-channel model, all available head channels (M1, M2, EOG1, EOG2) are first re-referenced to the average of the head montage. For recordings with limb EMG, the arm channels are converted to bipolar derivations.

In brief, saturated (clipped) samples are removed, and channels with abnormally low standard deviation (SD) are rejected before a high-pass filter (1 Hz) removes slow drifts, followed by a notch filter (50 or 60 Hz, depending on the mains frequency) to suppress power-line interference. The pipeline then performs iterative cleaning over the channel exhibiting the highest SD, and for amplitudes exceeding 400 µV, the offending samples and a surrounding window are flagged and masked. After each pass, the head channels are re-referenced to their group average for the 2-channel preprocessing, so high-amplitude transients do not bias the reference, and the procedure repeats until no violations remain. In the final phase, additional criteria remove residual presumed low-quality data. In recordings with limb EMG, periods of sustained high SD in the arm channels are excluded. Intervals with abnormally low root-mean-square (RMS) amplitude, indicative of signal attenuation or dropout, are also discarded. Finally, any surviving segment shorter than 3 s is removed, as such intervals are too short to support reliable analysis.

### U-Sleep training and fine-tuning

Automatic sleep staging used the U-Sleep architecture^25^, trained largely as described there with the following changes. During training of the 2-channel model, we applied a predefined channel-sampling scheme to enforce invariance to the specific electrode combination while preserving complementary EEG- and EOG-dominant views. From the two mastoid (M1, M2) and two EOG (EOG1, EOG2) electrodes, we formed a pool of nine derivations: the mastoid derivation M1-M2, the EOG derivation EOG1-EOG2, the four mastoid-to-EOG derivations (M1-EOG1, M1-EOG2, M2-EOG1, M2-EOG2), and the four electrode-to-head-average derivations. Each 17.5-min training sample paired two distinct derivations where the first (EEG view) was M1-M2 with probability 0.5 and otherwise drawn uniformly from the eight mixed derivations, and the second (EOG view) was drawn uniformly from the bipolar EOG and the eight mixed derivations, excluding the one already chosen.

A Base Model was first trained on a publicly available non-neurodegenerative cohort (Base Dataset) to learn general sleep architecture using 94%, 3%, and 3% of the subjects for training, validation, and testing, respectively. For the remaining datasets, all subjects were pooled and each was assigned to one of 10 folds, stratified by available recording type (PSG, Dual, Home). The Base Model was fine-tuned on the PACE-CBC Dataset (394 subjects, 443 recordings) using eight folds for training, one for validation, and one for testing. Repeating this across all 10-fold rotations yielded 10 model instances and unbiased predictions for every recording, since each subject is always scored by a model that never saw this subject in training or validation. Using the identical splits, we further fine-tuned the 10 instances on the home device recordings of the Dual Dataset (79 subjects, 84 recordings) to capture device-specific and electrode-placement-specific differences between polysomnography and home devices.

### Inference ensemble and rejection

At inference, the sampling scheme was treated as an ensemble, running the model independently over all 73 valid channel-pair combinations and aggregating their per-epoch, per-stage confidence scores into a single consensus hypnogram through three quality-weighted steps to make staging robust to noisy, NaN or detached electrodes by shifting weight to the remaining derivations.

1. **Global check**: For each pair, the NaN fraction for each channel is computed across the entire recording. If either channel exceeds 0.75, the pair is dropped from the ensemble.
2. **Local check**: In retained pairs, NaN gaps are interpolated for continuity, but any 30-s epoch with >5% NaNs is marked invalid for that pair and excluded from aggregation.
3. **Aggregation**: For each epoch, the softmax distributions over all valid pairs are averaged, and the stage is taken as the argmax of the resulting distribution. Epochs with no valid pair are labelled unknown.

### RWA estimation

We applied a rule-based algorithm to estimate the proportion of REM sleep without atonia during stage R using bipolar channels derived from the mastoid electrodes (M1-M2) and electrodes on the flexor digitorum superficialis muscles of the arms (FDS-EMG, bilateral pairs of electrodes). A combined ("Overall") estimate of RWA was computed as the union of the activity in the mastoid and both arm channels, meaning that activity occurring simultaneously in more than one channel contributes only once to the total proportion.

RWA was quantified with a rule-based approach. The algorithm applies automated arousal detection and distinct rules for temporal summation of activity rather than replicating the manual SINBAR 3s mini-epoch scheme, and it is not intended to reproduce the absolute RWA proportion of a human scorer. Before passing stage R epochs to the RWA estimator, an 80% stage R confidence threshold was applied as described previously^26^. Stage R was analysed in discrete periods, and recordings with fewer than 10 qualifying stage R epochs (5 min) were not scored. In brief, the detector takes the predicted REM epochs, evaluates a bipolar EMG-channel (mastoid-EMG, left or right FDS-EMG) and returns a binary RWA signal for every sample for the channel. Each channel was band-pass filtered with a zero-phase FIR filter (40-80 Hz, notch filtered during initial preprocessing). A period-specific baseline was defined as the 10^th^ percentile of the root-mean-square (RMS) amplitude over non-overlapping 30 s windows, and RWA activity was flagged wherever the RMS in a 0.5 s sliding window (0.24 s step) exceeded twice this baseline, and single-step gaps were bridged by morphological closing. Characteristics of arousals were then detected on the mastoid channel and removed from all activity masks. These presumed arousals were defined as an abrupt shift in the mastoid EEG-band signal, quantified by Hjorth mobility with the same windowing and thresholded at twice its 10^th^-percentile baseline, and with a duration adapting the AASM definition as lasting at least 3 s and bordered by at least 10 s of stable sleep. Sustained high-RMS bouts (≥15 s), taken to indicate wakefulness, were excluded in the same way. Because arousals were identified only from the mastoid but reflect global state, the flagged intervals were removed from both the mastoid and the arm results.

RWA was expressed as the fraction of approved REM duration (artefact-free signal within qualifying REM periods) containing detected RWA activity and computed separately for the mastoid-EMG and FDS-EMG channels and for their union.

### Determination of thresholds

RBD was classified using logistic regression on the automated RWA features. Subjects without a validated RBD label or with insufficient REM for feature estimation were excluded. Performance was estimated by leave-one-out cross-validation, refitting the full pipeline for each held-out subject, and the operating threshold was set at the maximum of Youden’s J on the leave-one-out predictions.

### Statistical analyses

Continuous variables are reported as mean (SD) or median [IQR] according to normality and categorical variables as counts (%). Staging agreement was quantified per recording by accuracy, Cohen’s kappa and stage-wise F1 with each recording weighted equally. Kappa was computed over the whole recording in the Base Dataset, where lights on and off annotations were not available, and over the lights-off period in the PACE-CBC Dataset. Differences in kappa and stage F1 between models were tested with two-sided Wilcoxon signed-rank tests. Agreement between manual and model-derived clinical sleep metrics was tested for each montage with two-sided paired t-tests on complete pairs and Holm-corrected.

RWA was compared across diagnostic groups with the Kruskal-Wallis test followed by Dunn’s test with Holm correction. Test-retest reliability of home RWA was assessed using the intraclass correlation coefficient. Across the three home nights, diagnostic yield was compared with Cochran’s Q, REM quantity with the Friedman test, and per-feature RWA with linear mixed models with a random intercept per subject with night 1 as the reference. Per-night classifier AUCs were compared with DeLong’s test, Holm-corrected across pairwise night comparisons.

Diagnostic discrimination used the area under the ROC curve with sensitivity and specificity at the Youden-optimal threshold. A two-sided alpha of 0.05 was used throughout. All analyses were performed in Python.

## Acknowledgements

We thank all study participants for their participation.

C.S. received funding from Aase and Ejnar Danielsen’s Foundation. A.J.T. received funding from the Jascha Foundation. P.B. received funding from The Independent Research Fund Denmark, The Riisfort Foundation, Parkinsonforeningen, the Lundbeck Foundation (R359-2020-2533, R491-2024-1966) and the Michael J. Fox Foundation (MJFF-022856).

M.S. received funding from the program "Netzwerke 2021", an initiative of the Ministry of Culture and Science of the State of North Rhine Westphalia, the Federal Ministry of Research, Technology and Space (BMFTR) under the funding code (FKZ): 01EO2107 and funding under the umbrella of the Partnership Fostering a European Research Area for Health (ERA4Health) (GA N° 101095426 of the EU Horizon Europe Research and Innovation Programme), and the European Research Council (ID 10116958).

The funders had no role in study design, data collection and analysis, the decision to publish or the preparation of the manuscript.

## Data availability

The Base Dataset is openly available from its original providers. The clinical datasets collected and analysed in this study are not publicly available owing to local ethics and data-protection regulations. Anonymised data in processed formats may be shared on reasonable request after obtaining the necessary ethical and data-sharing approvals.

